## Supplemental material for "Prospective validation of host transcriptomic biomarkers for pulmonary tuberculosis by real-time PCR"

**Appendix to:**

### Blood transcriptomic biomarkers for pulmonary tuberculosis: A prospective, multicentre, head-to-head validation by real-time PCR in a community setting

Simon C. Mendelsohn, Stanley Kimbung Mbandi, Andrew Fiore-Gartland, Adam Penn-Nicholson, Munyaradzi Musvosvi, Humphrey Mulenga, Michelle Fisher, Katie Hadley, Mzwandile Erasmus, Onke Nombida, Michèle Tameris, Gerhard Walzl, Kogieleum Naidoo, Gavin Churchyard, Mark Hatherill, Thomas J. Scriba, The Cross-sectional TB Cohort Study Team, and The CORTIS Study Team

#### Contents

**The CORTIS Study Team** 2
**The Cross-sectional TB Cohort Study Team** 4
**Supplemental Methods** 5
**Supplemental Results** 6
**Supplemental Tables** 7
**Supplemental Figures** 8
**References** 18
**STARD 2015 Checklist: An updated list of essential items for reporting diagnostic accuracy studies** 19
**Statistical Analysis Plan** 20

#### The CORTIS Study Team

*Centre for the AIDS Programme of Research in South Africa (CAPRISA), and MRC-CAPRISA HIV-TB Pathogenesis and Treatment Research Unit, Doris Duke Medical Research Institute, University of KwaZulu-Natal, Durban, South Africa.*
Bianca Bande
Thilagavathy Chinappa
Cara-Mia Corris
Thobelani Cwele
Celaphiwe Dlamini
Dhineshree Govender
Goodness Khanyisile Gumede
Nonhlanhla Zanele Elsie Gwamanda
Senzo Halti
Razia Hassan-Moosa
Senzo Ralph Hlathi
Zandile Patrica Jali
Lungile Khanyile
Bhavna Maharaj
Jabulisiwe Lethabo Maphanga
Nonhle Bridgette Maphanga
Siyabonga Mbatha
Atika Moosa
Nompumelelo Ngcobo
Ntombozuko Gloria Ntanjana
Sphelele Simo Nzimande
Nesri Padayatchi
Dirhona Ramjit
Thandiwe Yvonne Shezi
Zibuyile Phindile Penlee Sing
Chandrapharbha Singh
Philile Thembela
Londiwe Zaca
Mbali Ignatia Zulu

*Department of Molecular Microbiology, Washington University in St. Louis, St. Louis, MO, USA*
Shabaana A. Khader

*Division of Medical Microbiology, Department of Pathology, University of Cape Town, Cape Town, South Africa*
Slindile Mbhele
Mark P. Nicol
Judi van Heerden

*DST/NRF Centre of Excellence for Biomedical TB Research and SAMRC Centre for TB Research, Division of Molecular Biology and Human Genetics, Department of Biomedical Sciences, Faculty of Medicine and Health Sciences, Stellenbosch University, Cape Town, South Africa.*
Petri Ahlers
Roslyn Beukes
Maria Didloff
Marika Flinn
Bernadine Fransman
Andriëtte Hiemstra
Justine Khoury
Belinda Kriel
Jaftha Kruger
Andre Loxton
Elizna Maasdorp
Stephanus T. Malherbe
Onesisa Mpofu
Mpho Mtlali
Liesel Muller
Bronwyn Smith
Dorothy Solomons
Kim Stanley
Susanne Tonsing
Khayalethu Toto
Ayanda Tsamane
Susanne Tönsing

*South African Tuberculosis Vaccine Initiative, Institute of Infectious Disease and Molecular Medicine, Division of Immunology, Department of Pathology, University of Cape Town, South Africa.*
Charmaine Abrahams
Hadn Africa
Denis Arendsen
Nomfuneko Cynthia Batyi
Nicole Bilek
Natasja Botes
Samentra Braaf
Sivuyile Buhlungu
Alida Carstens
Balie Carstens
Nompumelelo Cetywayo
Yolundi Cloete
Lorraine Coetzee
Alessandro Companie
Ilse Davids
Marwou de Kock
Bongani Diamond
Palesa Dolo
Margareth Erasmus
Juanita Ferreira
Christal Ferus
Elizabeth Filander
Hennie Geldenhuys
Diann Gempies
Yolande Gregg
Rieyaat Hassiem
Roxane Herling
Yulandi Herselman
Chris Hikuam
Henry Issel
Ruwiyda Jansen
Lungisa Jaxa
Fabio Julies
Fazlin Kafaar
Masooda Kaskar
Sophie Keffers
Xoliswa Kelepu
Gloria Khomba
Sandra Kruger
Sunelza Lakay
Thelma Leopeng
Angelique Kany Kany Luabeya
Simbarashe Mabwe

Lauren Mactavie
Nomsitho Magawu
Lebohgang Makhete
Lebohang Makhethe
Faheema Meyer
Miriam Moses
Boitumelo Mosito
Angelique Mouton
Julia Noble
Nambitha Nqakala
Fajwa Opperman
Christel Petersen
Patiswa Plaatjie
Abe Pretorius
Rodney Raphela
Frances Ratangee
Maigan Ratangee
Susan Rossouw
Elisma Schoeman
Constance Schreuder
Alison September
Cashwin September
Justin Shenje
Marcia Steyn
Sonia Stryers
Liticia Swanepoel
Anne Swarts
Asma Toefy
Petrus Tyambetyu
Habibullah Valley
Linda van der Merwe
Elma van Rooyen
Ashley Veldsman
Helen Veldtsman
Kelvin Vollenhoven
Elaine Zimri

*TB Modelling Group, TB Centre, Centre for Mathematical Modelling of Infectious Diseases, Department of Infectious Disease Epidemiology, London School of Hygiene & Tropical Medicine, London, United Kingdom.*
Tom Sumner
Richard G. White

*The Aurum Institute, Johannesburg, Gauteng, South Africa.*
*Aurum Klerksdorp Site*
Tebogo Badimo
Kagiso Baepanye
Kesenogile Edna Baepanye
Tshepiso Baepanye
Ken Clarke
Marelize Collignon
Audrey Lebohang Dhlamini
Audrey Dlamini
Candice Eyre
Tebogo Feni
Moogo Fikizolo
Phinda Galane
Alia Gangat
Thelma Goliath
Craig Innes
Bonita Janse van Rensburg
Elba Janse van Rensburg
Olebogeng Jonkane
Boitumelo Sophy Kekana
Gomotsegang Virginia Khobedi
Marietjie King
Adrianne Kock
Ndlela Israel Kunene
Aneesa Lakhi
Nondumiso Langa
Hildah Ledwaba
Marillyn Luphoko
Immaculate Mabasa
Tshegofatso Dorah Mabe
Nkosinathi Charles Mabuza
Molly Majola
Mantai Makhetha
Blossom Makhubalo
Mpho Makoanyane
Vernon Malay
Shirley Malefo-Grootboom
Juanita Market
Selvy Matshego
Lungile Mbata
Nontsikelelo Mbipa
John Mdlulu
Tsiamo Mmotsa
Karabo Moche
Sylvester Modipa
Joseph Panie Moloko
Kabelo Molosi
Samuel Mopati
Palesa Moswegu
Primrose Mothaga
Dorothy Muller
Grace Nchwe
Nhlamulo Ndlovu
Maryna Nel
Lindiwe Nhlangulela
Tanya Nielson
Bantubonke Bertrum Ntamo
Lawerence Ntoahae
Tedrius Ntshauba
Thandiwe Papalagae
Pedro Pinho
Pearl Nomsa Sanyaka
Sharfuddin Sayed
Letlhogonolo Seabela
Raesibe Agnes Pearl Selepe
Melissa Neo Senne
Moeti Serake
Naydene Slabbert
Constance Takavamanya
Marthinette Taljaard
Maria Thlapi
Mugwena Thompo
Vincent Tshikovhi
Lebogang Isaac Tswaile
Amanda van Aswegen
Marietjie Zietsman

*Aurum Rustenburg Site*
Laudicia Tshenolo Bontsi

Obakeng Peter Booi
Mari Cathrin Botha
William Brumskine
Selemeng Matseliso Carol
Kgomotso Violet Chauke
Mooketsi Theophillius Cwaile
Isabella Johanna Davies
Emilia De Klerk
Blanchard Mbay Iyemosolo
James Michael Jeleni
Christian Mabika Kasongo
Sebaetseng Jeanette Kekana
Lucky Sipho Khoza
Gloria Keitumetse Kolobe
Lerato Julia Lekagane
Sheiley Christina Lekotloane
Ilze Jeanette Louw
Sarah Teboso Lusale
Perfect Tiisetso Maatjie
Kamogelo Fortunate Mabena
Johanna Thapelo Madikwe
Octavia Mahkosazana Madikwe
Rapontwana Letlhogonolo Maebana
Malobisa Sylvester Magwasha
Vutlhari-I-Vunhenha Fairlord Manzini
Isholedi Samuel Maroele
Omphile Petunia Masibi
July Rocky Mathabanzini
Tendamudzimu Ivan Mathode
Ellen Ditaba Matsane
Lungile Mbata
Nyasha Karen Mhandire
Thembisiwe Miga
Caroline Mkhokho
Neo Hilda Mkwalase
Nondzakazi Mnqonywa
Brenda Matshidiso Modisaotsile
Patricia Pakiso Mokgetsengoane
Kegomoditswe Magdeline Molatlhegi
Thuso Andrew Molefe
Motlatsi Evelyn Molotsi
Tebogo Edwin Montwedi
Boikanyo Dinah Monyemangene
Hellen Mokopi Mooketsi
Tshplpfelo Mapula Mosito
Ireen Lesebang Mosweu
Banyana Olga Motlagomang
Funeka Nomvula Mthembu
Themba Phakathi
Mapule Ozma Phatshwane
Victor Kgothatso Rameetse
Kelebogile Magdeline Segaetsho
Ni Ni Sein
Melissa Neo Senne
Sifiso Cornelius Shezi
Zona Sithetho
Bongiwe Stofile
Mando Mmakhora Thaba
Nosisa Charity Thandeka
Lethabo Collen Theko
Dimakatso Sylvia Tsagae

*Vaccine and Infectious Disease Division, Fred Hutchinson Cancer Research Center, Seattle, WA, USA.*
Bhavesh Borate
Eva Chung
Michelle Chung
Ellis Hughes
Alicia Sato
Steven Self

#### The Cross-sectional TB Cohort Study Team

*South African Tuberculosis Vaccine Initiative, Institute of Infectious Disease and Molecular Medicine, Division of Immunology, Department of Pathology, University of Cape Town, South Africa.*
Hadn Africa
Janelle Botes
Fatoumatta Darboe
Elizabeth Filander
Lebohang Makhethe
Sindile Matiwane
Melissa Murphy
Constance Schreuder
Marcia Steyn
Michele van Rooyen
Noncedo Xoyana

*Desmond Tutu HIV Centre, and Institute of Infectious Disease and Molecular Medicine (IDM), University of Cape Town, Cape Town, South Africa*
Carl Morrow
Robin Wood

#### Supplemental Methods

*Measurement of transcriptomic signatures*

The RISK11 transcriptomic signature was measured as previously described with pre-qualified TaqMan gene expression primer-probe assays.[1-3] The Roe1[4] (BATF2) and Roe3[5] signatures scores were calculated from the same Fluidigm 96.96 gene expression chip assay panels as the RISK11 signature (**Table S2**) by subtracting the geometric mean of four housekeeping gene raw cycle threshold (Ct) values from the average raw Ct of the genes of interest (**Table S1**).

A panel of 24 TaqMan gene expression primer-probe assays for the other seven signatures (**Table S3**)—Francisco2[6], Herberg2[7], Maertzdorf4[8], Penn-Nicholson6 (RISK6; **Table S4**)[9], Suliman4 (RISK4; **Table S5**)[10], Sweeney3[11], and Thompson5 (RESPONSE5; **Table S6**)[12]—were run in parallel on Fluidigm 192.24 gene expression chips. Primer-probe assays were qualified to ensure efficient amplification, linearity, and multiplexing capability, using methods similar to those described by Dominguez et al.[13] Briefly, we used five pre-amplified cDNA samples of varying concentration with a 12-point, two-fold dilution series. Each of the five dilution series were divided into eight segments of five consecutive dilutions and analysed for linearity in an iterative manner using the linear-least squares regression between the logarithm of the cDNA concentration and amplification cycle (Et; 40-Ct). An assay passed qualification if at least one segment within a dilution series met the following three criteria: (1) strong correlation (Pearson $r^2 \geq 0.99$), (2) linear slope (regression coefficient between 3.1–3.6), and (3) efficient amplification ($[10^{1/slope} - 1] = 90–110\%$).

*Quality control and analysis*

Fluidigm 192.24 gene expression chips were analysed using a locked-down R script (bitbucket.org/satvi/sixs) with quality control filters that assessed the integrity and reproducibility of each chip. The following parameters were applied for extracting Ct values: Linear (Derivative) baseline correction, Quality Threshold of 0.3, and Auto (Global) for Ct Threshold Method using Fluidigm software version 4.5.1. Chips with marked deviation in internal positive control sample primer-probe assay Ct values or RISK6 signature score from historical runs, with detection in the no-template (water) control or no-reverse-transcriptase (to detect amplification of genomic DNA ) control in primer-probes spanning exon-exon junctions, or with more than 20% failed reactions for a particular primer-probe assay, were repeated. Individual samples with more than 20% failed primer-probe reactions were classified as failed and no signature scores were computed. If less than 20% of primer-probe reactions failed for an individual sample, signature scores were computed where possible and signatures with missing primer-probe raw Ct values were deemed failed for that sample. Samples and primer-probe assays were run in singlet, and failed signature results for individual samples were assumed to follow a random distribution, thus not repeated, and excluded from analysis.

#### Supplemental Results

*Signature failure rate*

Although different pass rates may have biased comparison of performance between signature, the primary aim of the study was to compare individual signature performance to the WHO Target Product Profile (TPP) criteria benchmarks rather than against each other. Hence samples with failed signatures were still included in analysis to increase statistical power. Failure rate is also an indicator of signature robustness; an important criterion when selecting biomarkers for further development and clinical translation. The high failure rate of the Suliman4 signature was attributed to the GAS6 primer-probe assay, which performed well in assay qualification (**Table S3**) and validation in high yield and purity manually extracted PAXgene RNA samples. However, performance of the GAS6 assay was less robust in lower yield and quality RNA from automated robotic extraction. Due to the pair-wise ensemble structure of three up- and three down-regulated genes, RISK6 score can still be calculated even if one or more transcript is not detected due to failed PCR amplification.[9] As a result, RISK6 was resilient to single failed PCR reactions, with highest signature pass rate. Only HIV-uninfected participants with successful measurement of the RISK11 score were enrolled in the CORTIS-01 study, whereas HIV-infected participants were enrolled irrespective of RISK11 score in CORTIS-HR. This effectively excluded participants in CORTIS-01 with very low yield or purity RNA samples, resulting in a lower signature failure rates in CORTIS-01 as compared to CORTIS-HR.

#### Supplemental Tables

**Table S1.** Parsimonious transcriptomic signatures included in panel and signature score calculation

**Table S2.** TaqMan PCR primer-probe panel for Darboe11 (RISK11) Fluidigm 96.96 gene expression integrated fluidic circuit

**Table S3.** TaqMan PCR primer-probe panel for Fluidigm 192.24 gene expression integrated fluidic circuit and assay qualification

**Table S4.** Penn-Nicholson6 (RISK6) signature score calculation

**Table S5.** Suliman4 (RISK4) signature score calculation

**Table S6.** Thompson5 (RESPONSE5) signature score calculation

**Table S7.** Diagnostic performance of transcriptomic signatures in CTBC cohort

**Table S8.** Baseline characteristics of enrolled CORTIS-01 study cohort and tuberculosis endpoints

**Table S9.** Baseline characteristics of enrolled CORTIS-HR study cohort and tuberculosis endpoints

**Table S10.** Primary endpoint diagnostic performance of transcriptomic signatures in CORTIS-01 cohort

**Table S11.** Primary endpoint diagnostic performance of transcriptomic signatures in CORTIS-HR cohort

**Table S12.** Primary endpoint prognostic performance of transcriptomic signatures in CORTIS-01 cohort

**Table S13.** Primary endpoint prognostic performance of transcriptomic signatures in CORTIS-HR cohort

**Table S14.** Baseline characteristics of screened CORTIS-01 study participants enrolled in the respiratory pathobionts sub-study

**Table S15.** CTBC cohort signature scores and metadata

**Table S16.** CORTIS-01 sub-study cohort signature scores and metadata

**Table S17.** CORTIS-HR sub-study cohort signature scores and metadata

**Table S18.** CORTIS-01 Respiratory Pathogens sub-study cohort signature scores and metadata

#### Supplemental Figures

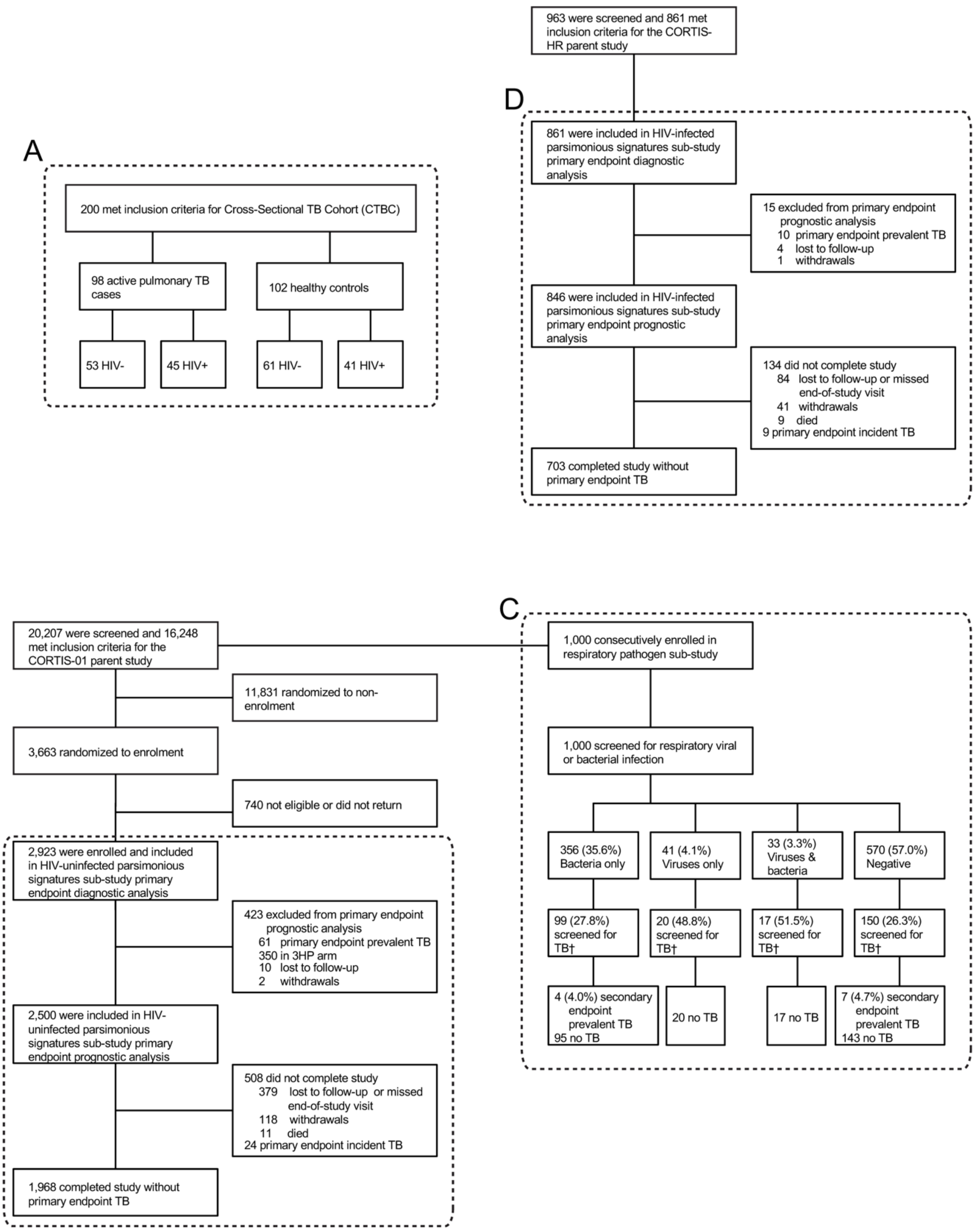


**Figures S1: Study flow diagram**

(A) HIV-infected and HIV-uninfected participants with and without TB were recruited into the Cross-Sectional TB Cohort (CTBC) case-control study. HIV-uninfected participants were recruited from the CORTIS-01 parent study and co-enrolled into the (B) HIV-uninfected parsimonious signatures sub-study and (C) respiratory pathobionts sub-study. HIV-infected participants were recruited from the CORTIS-HR parent study and co-enrolled into the (D) HIV-infected parsimonious signatures sub-study. For the CTBC study and respiratory pathobionts sub-study, TB disease was defined by a single sputum sample positive for *Mtb* on either Xpert MTB/RIF and/or liquid culture at enrolment. The coprimary endpoints in the CORTIS studies were baseline prevalent TB disease and incident disease through 15 months follow-up confirmed by a positive Xpert MTB/RIF, Xpert Ultra, or MGIT culture, on two or more separate sputum samples collected within any 30-day period. The secondary endpoint was microbiologically-confirmed TB disease on at least one sputum sample.

†Out of 1000 participants enrolled in the respiratory pathobionts cohort, only 286 (28.6%) participants were co-enrolled in the CORTIS-01 parent study and investigated for TB at enrolment; 11/286 (3.8%) had prevalent TB confirmed by *Mycobacterium tuberculosis* liquid culture and/or Xpert MTB/RIF.

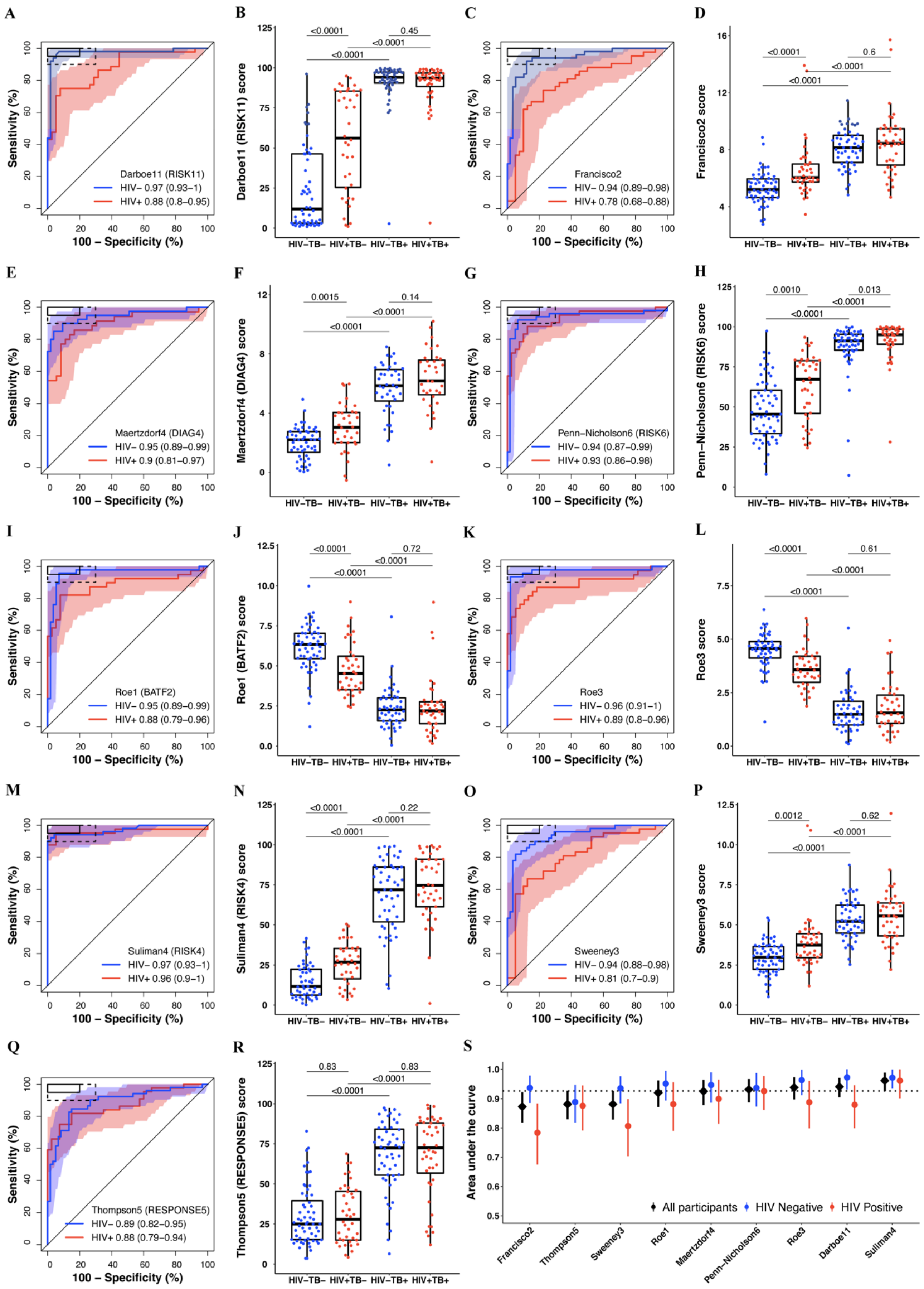


**Figures S2: Parsimonious signatures diagnostic performance and signature score distributions in the Cross-sectional Tuberculosis Cohort (CTBC) study**

Paired receiver operating characteristic (ROC) curves and box-and-whisker plots for the (A-B) Darboe11 (RISK11), (C-D) Francisco2, (E-F) Maertzdorf4 (DIAG4), (G-H) Penn-Nicholson6 (RISK6), (I-J) Roe1 (BATF2), (K-L) Roe3, (M-N) Suliman4 (RISK4), (O-P) Sweeney3, and (Q-R) Thompson5 (RESPONSE5) signatures in the Cross-sectional Tuberculosis Cohort (CTBC) study.

The ROC curves depict diagnostic performance (area under the curve, AUC, with 95% CI) of the parsimonious signatures for diagnosing TB, stratified by HIV status. The shaded areas represent the 95% CIs. The solid box depicts the optimal criteria (95% sensitivity and 80% specificity) and the dashed box depicts the minimal criteria (90% sensitivity and 70% specificity) set out in the WHO Target Product Profile for a triage test.[14]

The box-and-whisker plots depict signature score distribution by HIV (HIV+/HIV-) and prevalent TB disease (TB+/TB-) status. Each dot represents a participant. p values for comparison of median signature scores between groups in box-and-whisker plots were calculated with the Mann-Whitney *U* test and corrected for multiple comparisons by use of the Benjamini-Hochberg Procedure.[15] Boxes depict the IQR, the midline represents the median, and the whiskers indicate the IQR ± (1.5 × IQR).

(S) Summary of signature diagnostic performance in order of AUC estimates in all participants. The diagnostic AUC estimates in HIV-infected and HIV-uninfected participant sub-groups are also shown. The midline indicates the AUC estimate, the error bars indicate the 95% CIs, and the black dotted line indicates the lower bound of the 95% CI for the best performing signature for all participants.

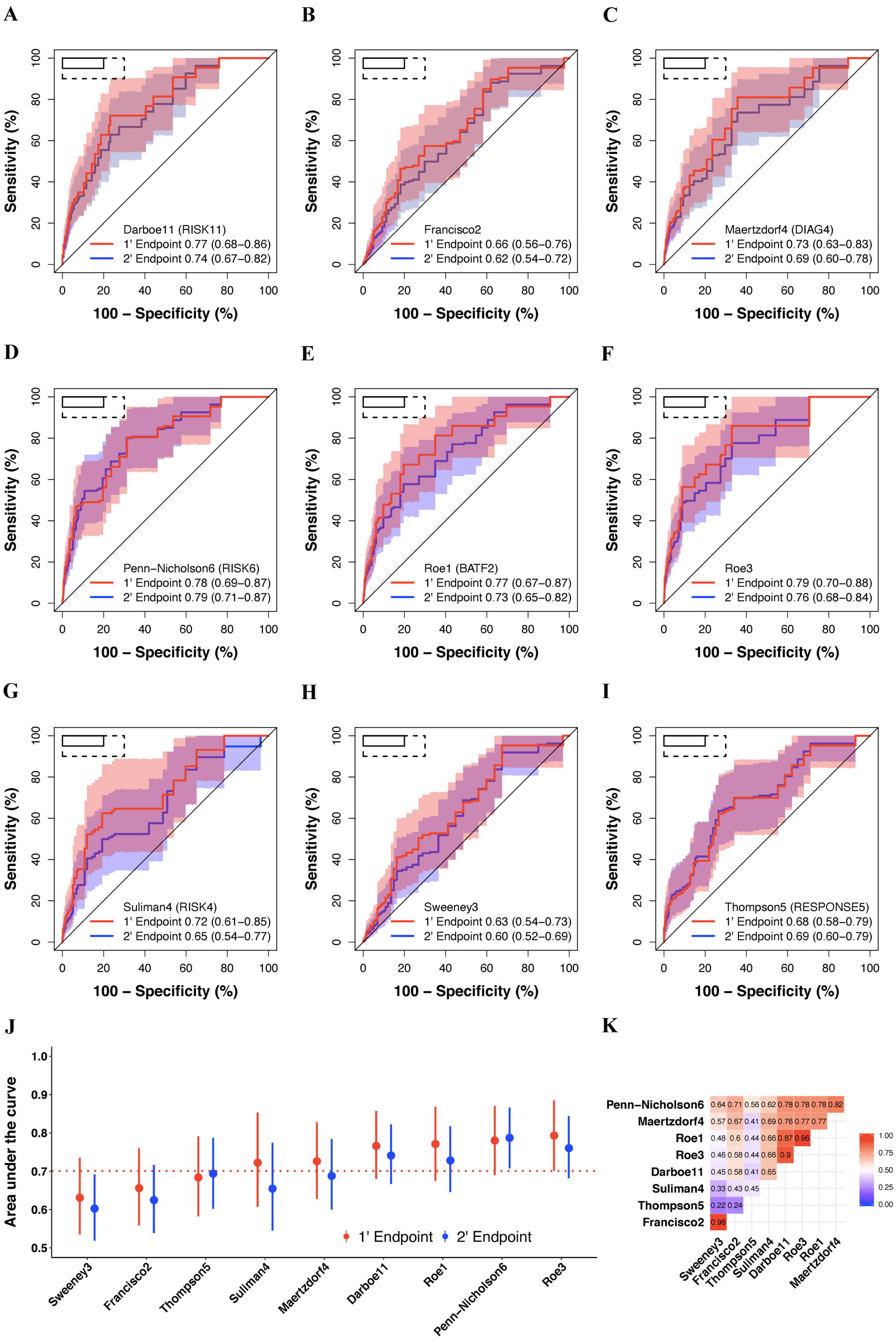


**Figures S3: Parsimonious signatures diagnostic performance for primary versus secondary endpoint TB in people without HIV**

Receiver operating characteristic (ROC) curves depicting primary ('1) versus secondary ('2) endpoint diagnostic performance (area under the curve, AUC, with 95% CI) of the (A) Darboe11 (RISK11), (B) Francisco2, (C) Maertzdorf4 (DIAG4), (D) Penn-Nicholson6 (RISK6), (E) Roe1 (BATF2), (F) Roe3, (G) Suliman4 (RISK4), (H) Sweeney3, and (I) Thompson5 (RESPONSE5) signatures in the CORTIS-01 study of people without HIV. The shaded areas represent the 95% CIs. The solid box depicts the optimal criteria (95% sensitivity and 80% specificity) and the dashed box depicts the minimal criteria (90% sensitivity and 70% specificity) set out in the WHO Target Product Profile for a triage test.[14]

(J) Summary of signature diagnostic performance in order of primary endpoint AUC estimates. The diagnostic AUC estimates for the secondary endpoint are also shown. The midline indicates the AUC estimate, the error bars indicate the 95% CIs, and the red dotted line indicates the lower bound of the 95% CI for the best performing signature for the primary endpoint.

(K) Signature score correlation matrix with the Spearman rank-order correlation coefficients. Roe1 and Roe3 signatures were multiplied by -1 to obtain a positive correlation for all signatures.

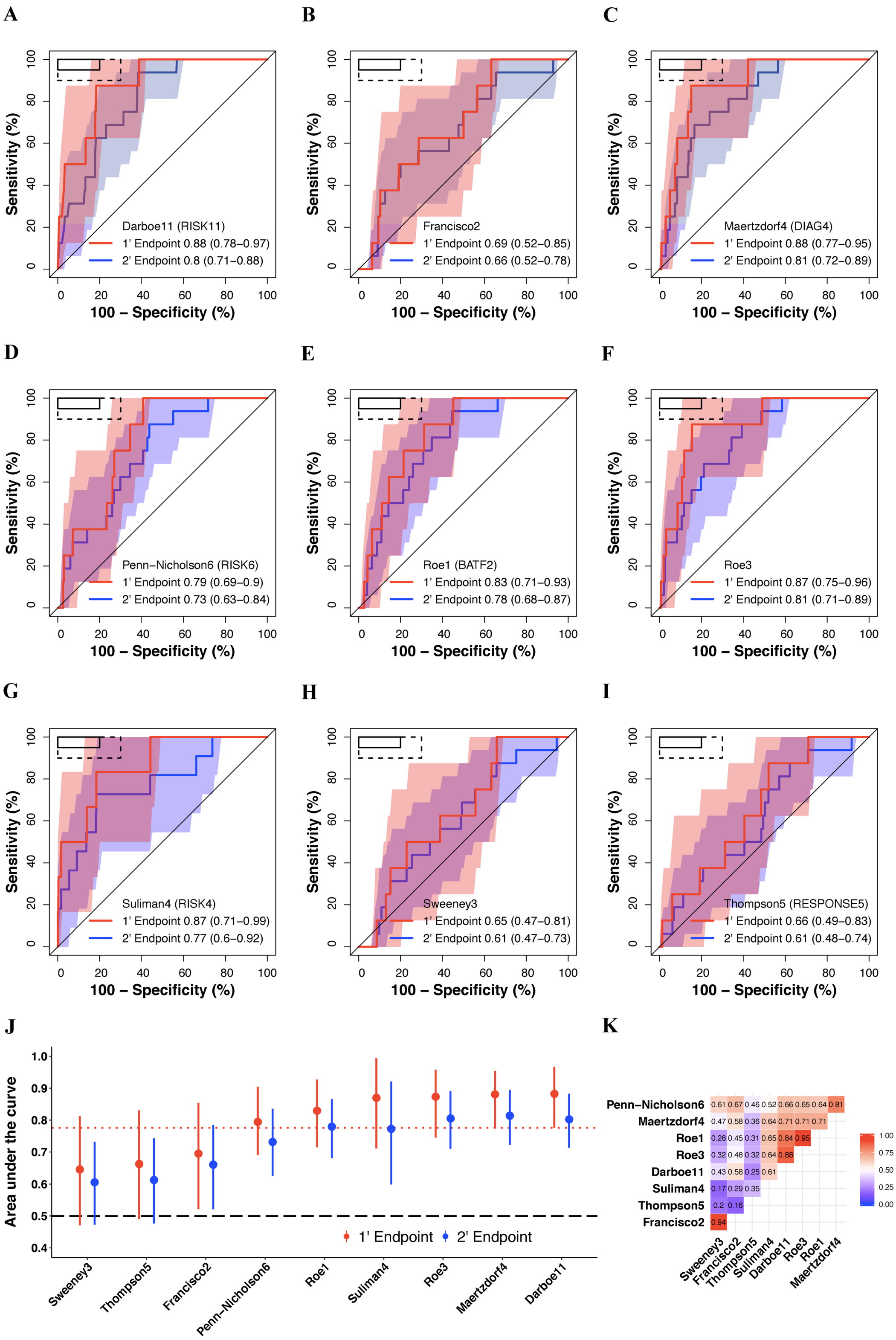


**Figures S4: Parsimonious signatures diagnostic performance for primary versus secondary endpoint TB in people living with HIV**

Receiver operating characteristic (ROC) curves depicting primary (‘1) versus secondary (‘2) endpoint diagnostic performance (area under the curve, AUC, with 95% CI) of the (A) Darboe11 (RISK11), (B) Francisco2, (C) Maertzdorf4 (DIAG4), (D) Penn-Nicholson6 (RISK6), (E) Roe1 (BATF2), (F) Roe3, (G) Suliman4 (RISK4), (H) Sweeney3, and (I) Thompson5 (RESPONSE5) signatures in the CORTIS-HR study of people living with HIV. The shaded areas represent the 95% CIs. The solid box depicts the optimal criteria (95% sensitivity and 80% specificity) and the dashed box depicts the minimal criteria (90% sensitivity and 70% specificity) set out in the WHO Target Product Profile for a triage test.[14]

(J) Summary of signature diagnostic performance in order of primary endpoint AUC estimates. The diagnostic AUC estimates for the secondary endpoint are also shown. The midline indicates the AUC estimate, the error bars indicate the 95% CIs, and the red dotted line indicates the lower bound of the 95% CI for the best performing signature for the primary endpoint. The black dashed line indicates an AUC cut-off of 0.5.

(K) Signature score correlation matrix with the Spearman rank-order correlation coefficients. Roe1 and Roe3 signatures were multiplied by -1 to obtain a positive correlation for all signatures.

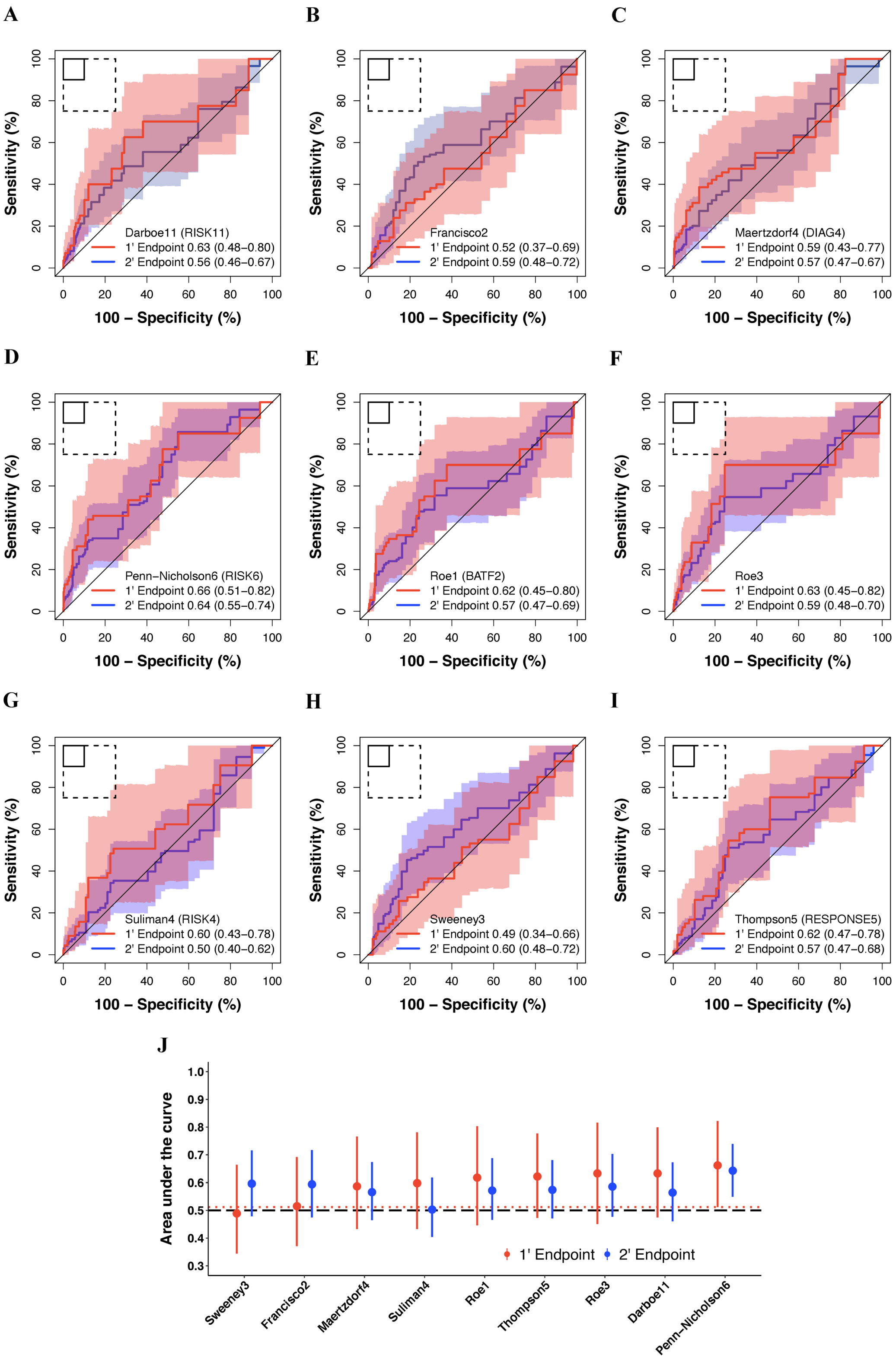


**Figures S5: Parsimonious signatures prognostic performance for primary versus secondary endpoint TB through 15-months follow-up in people without HIV**

Receiver operating characteristic (ROC) curves depicting primary (‘1) versus secondary (‘2) endpoint prognostic performance (area under the curve, AUC, with 95% CI) of the (A) Darboe11 (RISK11), (B) Francisco2, (C) Maertzdorf4 (DIAG4), (D) Penn-Nicholson6 (RISK6), (E) Roe1 (BATF2), (F) Roe3, (G) Suliman4 (RISK4), (H) Sweeney3, and (I) Thompson5 (RESPONSE5) signatures through 15-months follow-up in the CORTIS-01 study of people without HIV. The shaded areas represent the 95% CIs. The solid box depicts the optimal criteria (90% sensitivity and 90% specificity) and the dashed box depicts the minimal criteria (75% sensitivity and 75% specificity) set out in the WHO Target Product Profile for an incipient TB test.[16]

(J) Summary of signature prognostic performance in order of primary endpoint AUC estimates. The prognostic AUC estimates for the secondary endpoint are also shown. The midline indicates the AUC estimate, the error bars indicate the 95% CIs, the red dotted line indicates the lower bound of the 95% CI for the best performing signature for the primary endpoint, and the black dashed line indicates an AUC cut-off of 0.5.

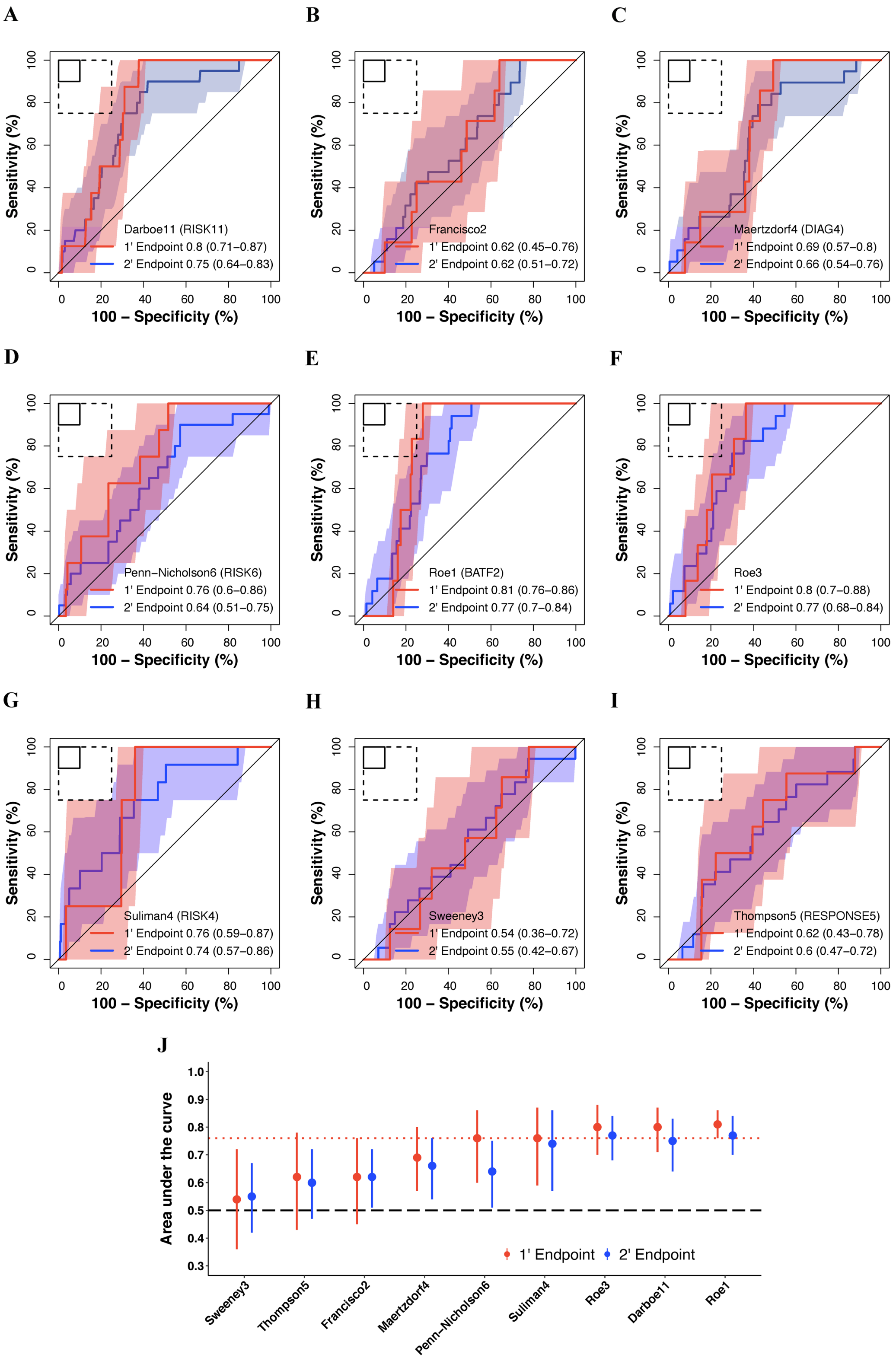


**Figures S6: Parsimonious signatures prognostic performance for primary versus secondary endpoint TB through 15-months follow-up in people living with HIV**

Receiver operating characteristic (ROC) curves depicting primary ('1) versus secondary ('2) endpoint prognostic performance (area under the curve, AUC, with 95% CI) of the (A) Darboe11 (RISK11), (B) Francisco2, (C) Maertzdorf4 (DIAG4), (D) Penn-Nicholson6 (RISK6), (E) Roe1 (BATF2), (F) Roe3, (G) Suliman4 (RISK4), (H) Sweeney3, and (I) Thompson5 (RESPONSE5) signatures through 15-months follow-up in the CORTIS-HR study of people living with HIV. The shaded areas represent the 95% CIs. The solid box depicts the optimal criteria (90% sensitivity and 90% specificity) and the dashed box depicts the minimal criteria (75% sensitivity and 75% specificity) set out in the WHO Target Product Profile for an incipient TB test.[16]

(J) Summary of signature prognostic performance in order of primary endpoint AUC estimates. The prognostic AUC estimates for the secondary endpoint are also shown. The midline indicates the AUC estimate, the error bars indicate the 95% CIs, the red dotted line indicates the lower bound of the 95% CI for the best performing signature for the primary endpoint, and the black dashed line indicates an AUC cut-off of 0.5.

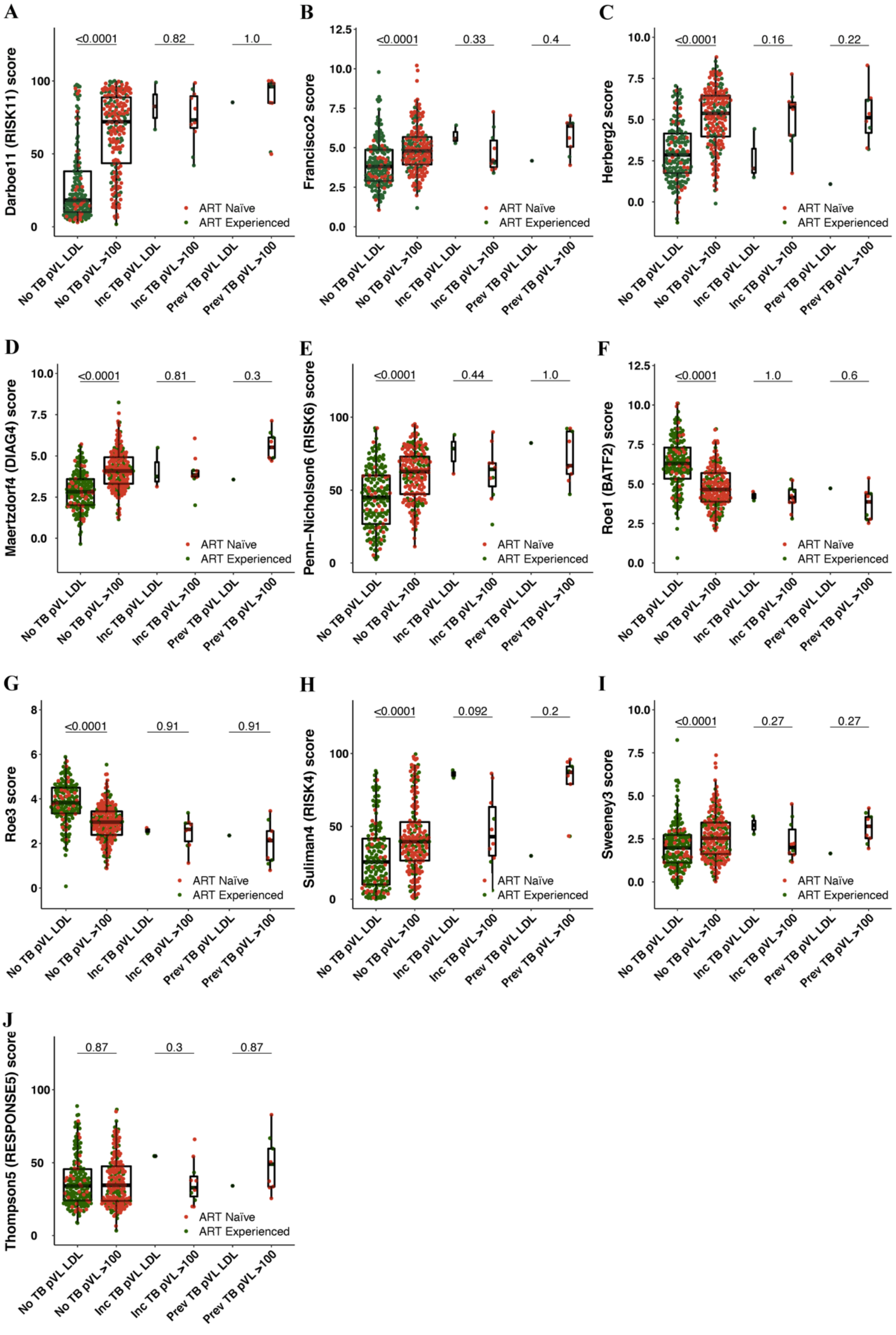


**Figures S7: Parsimonious signatures score distribution by HIV plasma viral load**

Box-and-whisker plots depicting distributions of the (A) Darboe11 (RISK11), (B) Francisco2, (C) Herberg2, (D) Maertzdorf4 (DIAG4), (E) Penn-Nicholson6 (RISK6), (F) Roe1 (BATF2), (G) Roe3, (H) Suliman4 (RISK4), (I) Sweeney3, and (J) Thompson5 (RESPONSE5) signature scores measured at baseline in the CORTIS-HR study stratified by HIV plasma viral load (copies per mL) and TB status. Prevalent (Prev) and incident (Inc) TB comprised all microbiologically confirmed secondary endpoint cases (i.e. TB confirmed on at least one sputum sample). HIV plasma viral load (pVL) is stratified into two groups: lower than the detectable limit (LDL) of 100 copies per mL and greater than 100 copies per mL (>100).

Each dot represents a participant. p values for comparison of median signature scores between groups in box-and-whisker plots were calculated with the Mann-Whitney *U* test and corrected for multiple comparisons by use of the Benjamini-Hochberg Procedure.[15] Boxes depict the IQR, the midline represents the median, and the whiskers indicate the IQR ± (1.5 × IQR).

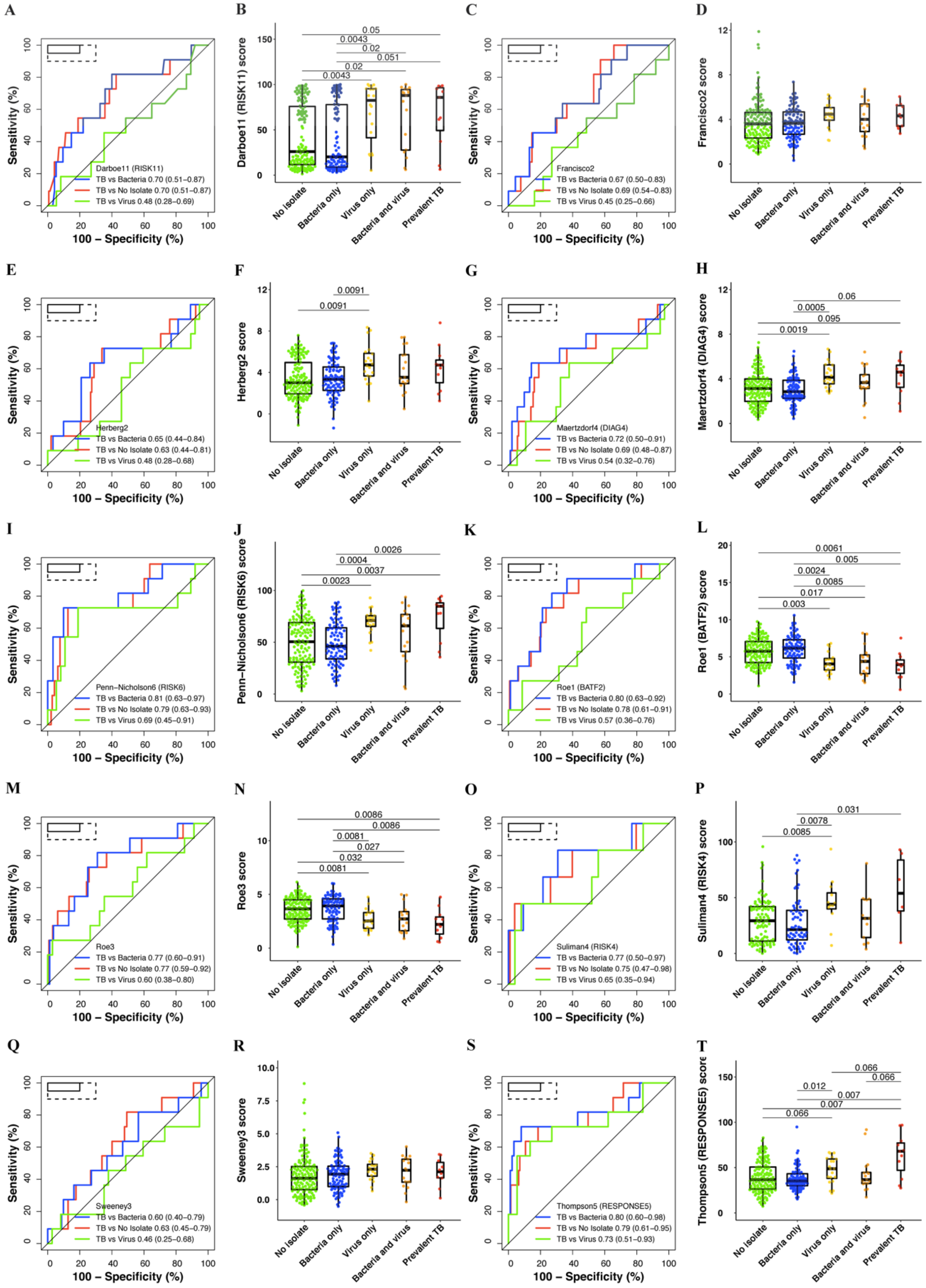


**Figures S8: Parsimonious signatures performance for differentiating participants with TB from those viral or bacterial upper respiratory tract pathobionts, and those without any pathobionts**

Paired receiver operating characteristic (ROC) curves and box-and-whisker plots for the (A-B) Darboe11 (RISK11), (C-D) Francisco2, (E-F) Herberg2, (G-H) Maertzdorf4 (DIAG4), (I-J) Penn-Nicholson6 (RISK6), (K-L) Roe1 (BATF2), (M-N) Roe3, (O-P) Suliman4 (RISK4), (Q-R)

Sweeney3, and (S-T) Thompson5 (RESPONSE5) signatures in all participants randomised in the CORTIS-01 study and co-enrolled in the respiratory pathobionts sub-study. Participants who were not randomised in the CORTIS-01 studied, and thus not investigated for TB, are not included.

The box-and-whisker plots depict signature score distribution in participants with no upper respiratory pathobionts (n=143), bacterial upper respiratory pathobionts only (n=95), viral upper respiratory pathobionts only(n=20), both viral and bacterial upper respiratory pathobionts (n=17), and *Mycobacterium tuberculosis* (n=11) detected on GeneXpert MTB/RIF or MGIT culture (microbiologically-confirmed secondary endpoint prevalent TB; i.e. TB confirmed on at least one sputum sample). p values for comparison of median signature scores between groups in box-and-whisker plots were calculated with the Mann-Whitney *U* test and corrected for multiple comparisons by use of the Benjamini-Hochberg Procedure.[15] Only p-values below 0.1 are shown. Each dot represents a participant. Boxes depict the IQR, the midline represents the median, and the whiskers indicate the IQR ± (1.5 × IQR).

The ROC curves depict performance (area under the curve, AUC, with 95% CI) of the parsimonious signatures in differentiating between participants with prevalent TB (n=11) and participants with viral upper respiratory pathobionts (n=37), participants with bacterial upper respiratory pathobionts only (n=95), and participants with no pathobionts (n=143). Participants with both viral and bacterial pathobionts (n=17) were included in the group with viral pathobionts only (n=20) as the presence of bacterial pathobionts did not appear to affect signature scores (Figure 5).

The solid box depicts the optimal criteria (95% sensitivity and 80% specificity) and the dashed box depicts the minimal criteria (90% sensitivity and 70% specificity) set out in the WHO Target Product Profile for a triage test.[14]

#### STARD 2015 Checklist: An updated list of essential items for reporting diagnostic accuracy studies

| Section & Topic | No | Item | Reported on page |
|---|---|---|---|
| **TITLE OR ABSTRACT** | | | |
| | **1** | Identification as a study of diagnostic accuracy using at least one measure of accuracy (such as sensitivity, specificity, predictive values, or AUC) | **p2** |
| **ABSTRACT** | | | |
| | **2** | Structured summary of study design, methods, results, and conclusions (for specific guidance, see STARD for Abstracts) | **p2** |
| **INTRODUCTION** | | | |
| | **3** | Scientific and clinical background, including the intended use and clinical role of the index test | **p3-4** |
| | **4** | Study objectives and hypotheses | **p4** |
| **METHODS** | | | |
| *Study design* | **5** | Whether data collection was planned before the index test and reference standard were performed (prospective study) or after (retrospective study) | **p5** |
| *Participants* | **6** | Eligibility criteria | **p5** |
| | **7** | On what basis potentially eligible participants were identified (such as symptoms, results from previous tests, inclusion in registry) | **p5-6** |
| | **8** | Where and when potentially eligible participants were identified (setting, location and dates) | **p5-6** |
| | **9** | Whether participants formed a consecutive, random or convenience series | **p5-6** |
| *Test methods* | **10a** | Index test, in sufficient detail to allow replication | **p7** |
| | **10b** | Reference standard, in sufficient detail to allow replication | **p8** |
| | **11** | Rationale for choosing the reference standard (if alternatives exist) | **N/A** |
| | **12a** | Definition of and rationale for test positivity cut-offs or result categories of the index test, distinguishing pre-specified from exploratory | **p9** |
| | **12b** | Definition of and rationale for test positivity cut-offs or result categories of the reference standard, distinguishing pre-specified from exploratory | **N/A** |
| | **13a** | Whether clinical information and reference standard results were available to the performers/readers of the index test | **p9** |
| | **13b** | Whether clinical information and index test results were available to the assessors of the reference standard | **p9** |
| *Analysis* | **14** | Methods for estimating or comparing measures of diagnostic accuracy | **p8-9** |
| | **15** | How indeterminate index test or reference standard results were handled | **Appendix p5-6** |
| | **16** | How missing data on the index test and reference standard were handled | **Appendix p5-6** |
| | **17** | Any analyses of variability in diagnostic accuracy, distinguishing pre-specified from exploratory | **N/A** |
| | **18** | Intended sample size and how it was determined | **p8** |
| **RESULTS** | | | |
| *Participants* | **19** | Flow of participants, using a diagram | **Figure S1** |
| | **20** | Baseline demographic and clinical characteristics of participants | **Tables S8, S9, and S14** |
| | **21a** | Distribution of severity of disease in those with the target condition | **N/A** |
| | **21b** | Distribution of alternative diagnoses in those without the target condition | **Figure S1C** |
| | **22** | Time interval and any clinical interventions between index test and reference standard | **p5-7** |
| *Test results* | **23** | Cross tabulation of the index test results (or their distribution) by the results of the reference standard | **Tables S7, S10-S13** |
| | **24** | Estimates of diagnostic accuracy and their precision (such as 95% confidence intervals) | **Tables S7, S10-S13** |
| | **25** | Any adverse events from performing the index test or the reference standard | **N/A** |
| **DISCUSSION** | | | |
| | **26** | Study limitations, including sources of potential bias, statistical uncertainty, and generalisability | **p19** |
| | **27** | Implications for practice, including the intended use and clinical role of the index test | **p19** |
| **OTHER INFORMATION** | | | |
| | **28** | Registration number and name of registry | **N/A** |
| | **29** | Where the full study protocol can be accessed | **Appendix p20** |
| | **30** | Sources of funding and other support; role of funders | **p10, 20-21** |

**Source:** Bossuyt PM, Reitsma JB, Bruns DE, Gatsonis CA, Glasziou PP, Irwig L, et al. STARD 2015: an updated list of essential items for reporting diagnostic accuracy studies. BMJ (Clinical Research Ed). 2015;351:h5527. doi: 10.1136/bmj.h5527.

### Statistical Analysis Plan:

#### Evaluation of parsimonious host-blood tuberculosis transcriptomic signatures in HIV-infected and HIV-uninfected individuals

##### *A sub-study of the CORTIS-01 and CORTIS-HR trials*

| | |
|---|---|
| **Description:** | This document describes the statistical analysis plan to be performed on data obtained from the above-named sub-study of the CORTIS-01 and CORTIS-HR studies and is to be read in conjunction with the approved CORTIS-01 study protocol version 2.0 (01 August 2017) and SAP version 3.0 (24 October 2019), and CORTIS-HR study protocol version 1.0 (26th August 2016) and SAP version 1.0 (11 December 2019). |

**Authors:**

Simon C Mendelsohn[1], MBChB

Andrew Fiore-Gartland[2], Ph.D.

[1]South African Tuberculosis Vaccine Initiative, Department of Pathology and Institute of Infectious Diseases and Molecular Medicine, University of Cape Town

[2]Statistical Center for HIV/AIDS Research and Prevention, Vaccine and Infectious Disease Division, Fred Hutchinson Cancer Research Center

**Date:** 08 January 2020

**Version:** v 1.0

| | |
|---|---|
| **Document:** | Statistical Analysis Plan: Evaluation of parsimonious host-blood tuberculosis transcriptomic signatures in HIV-infected and HIV-uninfected individuals.<br>*A sub-study of the CORTIS-01 and CORTIS-HR trials* |
| **SAP Version:** | 1.0 |
| **Version Date:** | 08 January 2020 |
| **Authorised By:** | Prof Mark Hatherill, MBChB<br>CORTIS-01 and CORTIS-HR Principal Investigator<br>South African Tuberculosis Vaccine Initiative, University of Cape Town<br><br>Sign: ________________________________<br><br>Date: ________________________________<br><br>Dr Andrew Fiore-Gartland, PhD<br>CORTIS-01 and CORTIS-HR Lead Statistician<br>Vaccine and Infectious Disease Division, Fred Hutchinson Cancer Research Center<br><br>Sign: ________________________________<br><br>Date: ________________________________ |

### Table of Contents

*1 List of Abbreviations* 5

*2 Statistical Analysis Plan Overview* 6

**2.1 Aims** 7

2.1.1 Primary aims 7

2.1.2 Secondary aims 7

**2.2 Study Endpoints** 7

2.2.1 Two-sample endpoint definition 7

2.2.1 One and two-sample endpoint definition 7

**2.3 TB disease endpoint adjudication algorithm and censoring** 7

**2.4 Study design** 8

**2.5 Measurement of parsimonious host-blood TB transcriptomic signature scores** 8

**2.6 Projected enrolment, case accrual and power** 9

**2.7 Analysis populations and weighting** 9

2.7.1 CORTIS-01 9

2.7.1 CORTIS-HR 10

*3 Statistical Considerations* 10

**3.1 General principles** 10

**3.2 Missing data** 10

**3.3 Responsibility** 10

**3.4 Blinding** 10

**3.5 Data storage** 10

*4 Statistical Methods* 10

**4.1 Signature performance analysis** 10

**4.2 Primary aim 1: Signature diagnostic performance** 11

**4.3 Primary aim 2: Signature predictive performance** 11

**4.4 Confidence intervals** 12

**4.5 Secondary aimS: Comparison of signature performance** 12

**4.6 Exploratory sub-group analyses** 12

4.6.1 CORTIS-01 and CORTIS-HR 13

4.6.2 CORTIS-HR only 13

*5 Tables* 14

**5.1 Primary aim 1: Signature diagnostic performance at enrolment (BINARY ANALYSIS) IN HIV-infected (CORTIS-HR) / HIV-uninfected (CORTIS-01) cohort** 14

**5.2 Primary Aim 2: Signature predictive performance for identification of TB disease over a 15-month period, stratified by the time interval to disease (Time-dependent analysis) in HIV-infected (CORTIS-HR) / HIV-uninfected (CORTIS-01) cohort ____ 16**

***6 Figures ____ 18***

**6.1 Test score distribution: violin/box-and-scatter plots. Plots will be weighted for CORTIS-01 (section 2.7.1) and unweighted for CORTIS-HR (section 2.7.2). ____ 18**

**6.2 Test score correlations: correlation matrix of signature scores versus signature scores and demographic variables (spearman *rho* correlation coefficient). Plots will be weighted for CORTIS-01 (section 2.7.1) and unweighted for CORTIS-HR (section 2.7.2). ____ 18**

**6.3 Test performance: ROC curves (sensitivity versus 100-specificity) with relevant WHO TPP criteria indicated ____ 18**

**6.4 Test accuracy: sensitivity/specificity versus test score threshold plots (x-axis: score threshold, y-axis: sensitivity or specificity) ____ 18**

**6.5 Time dependent analysis: AUC versus time. Sensitivity, specificity, NPV and PPV (at different thresholds) versus time, over 15-month follow-up ____ 18**

***7 Appendix ____ 19***

**7.1 WHO diagnostic, triage, and predictive TB test performance target product profile ____ 19**

***8 References ____ 20***

### 1 LIST OF ABBREVIATIONS

| | |
|---|---|
| AFB | Acid-fast bacilli |
| BMI | Body mass index |
| cDNA | Complementary DNA |
| CI | Confidence interval |
| CIR | Cumulative incidence ratio |
| COR | Correlate of risk |
| CRA | Clinical research associate |
| CRF | Case report form |
| eCRF | Electronic CRFs |
| GCP | Good clinical practice |
| $H_0$ | Null hypothesis |
| HIV | Human immunodeficiency virus |
| IEC | Independent ethics committee |
| IGRA | Interferon gamma release assay |
| INH | Isoniazid |
| IPT | Isoniazid preventive therapy |
| LAM | Lipoarabinomannan |
| LTBI | Latent tuberculosis infection |
| MDR-TB | Multi-drug resistant tuberculosis |
| MGIT | Mycobacteria growth indicator tube |
| mRNA | Messenger RNA |
| *Mtb* | *Mycobacterium tuberculosis* |
| OR | Odds ratio |
| NNS | Number needed to screen (to detect one case) |
| NPV | Negative predictive value |
| PI | Principal investigator |
| PPV | Positive predictive value |
| QFT | QuantiFERON |
| RR | Relative risk |
| $RR_{COR}(15)$ | Relative risk for TB disease over 15 months |
| SA | South Africa |
| SATVI | South African Tuberculosis Vaccine Initiative |
| TB | Tuberculosis |
| TCD | Triclinium Clinical Development |
| TST | Tuberculin skin test |
| WHO | World Health Organization |

### 2 STATISTICAL ANALYSIS PLAN OVERVIEW

This statistical analysis plan (SAP) is for a sub-study of the CORTIS-01 and CORTIS-HR trails and is to be read in conjunction with the approved CORTIS-01 study protocol version 2.0 (01 August 2017) and SAP version 3.0 (24 October 2019), and CORTIS-HR study protocol version 1.0 (26th August 2016) and SAP version 1.0 (11 December 2019).

There is a need for earlier TB case identification, using novel non-sputum based diagnostics, linked to more effective preventive and curative strategies (World Health Organization, 2015). A blood-based triage test that allows targeted investigation for active and sub-clinical TB disease, including asymptomatic individuals at highest risk of progression from latency to disease, could shorten the time to TB treatment, or even prevent disease before symptoms emerge. The tuberculin skin test (TST) and interferon gamma release assay (IGRA) have poor specificity for incident TB disease in endemic populations, including HIV infected people (Auguste, *BMC Infect Dis*, 2017).

We have previously developed a highly specific predictive correlate of risk (COR) to identify healthy, HIV uninfected, South African adults at high risk of active TB disease (Zak, *Lancet*, 2016). This validated COR, based on mRNA expression signatures in blood, prospectively discriminates between TB cases and healthy controls among HIV uninfected persons. Based on published microarray case-control datasets, the COR has 87% diagnostic sensitivity and 97% specificity for prevalent TB disease in HIV uninfected South African adults (Zak, *Lancet*, 2016); and in two nested case-control studies, also among HIV uninfected persons, the COR has 70% predictive sensitivity and 84% specificity for incident TB disease occurring within one year of sampling (Penn-Nicholson, *S Afr Med J*, 2016). This PCR-based mRNA COR signature has been refined to 11-genes (RISK11) with equivalent diagnostic performance (Darboe, *Tuberculosis*, 2018).

Although relatively parsimonious, this signature is not ideal for a point-of-care (POC) device because of its size. Several concise mRNA signatures have recently been developed which are translatable into a POC test device (Gupta, *bioRxiv*, 2019; Warsinske, *PLoS Med*, 2019). A POC device could be employed for test-and-treat strategies in the community and clinic, both to detect early (and asymptomatic) TB disease for curative treatment and to identify *M.tb* infected individuals at high risk of progression to active disease for targeted short-course preventive therapy.

The aim of this sub-study is to evaluate and compare the performance of parsimonious host-blood TB transcriptomic signatures to identify prevalent TB disease and predict incident TB disease in HIV-infected and HIV-uninfected adults.

#### 2.1 AIMS

##### *2.1.1 PRIMARY AIMS*

2.1.1.1 **Primary Aim 1:** Estimate whether parsimonious host-blood TB transcriptomic signatures differentiate HIV-infected (CORTIS-HR) and HIV-uninfected (CORTIS-01) persons with prevalent TB disease from those without prevalent TB disease.

2.1.1.2 **Primary Aim 2:** Estimate whether parsimonious host-blood TB transcriptomic signatures differentiate HIV-infected (CORTIS-HR) and HIV-uninfected (CORTIS-01) persons at high risk for incident TB disease from those at low risk for incident TB disease.

##### *2.1.2 SECONDARY AIMS*

2.1.2.1 **Secondary Aim 1:** Compare parsimonious host-blood TB transcriptomic signatures diagnostic performance in HIV-infected (CORTIS-HR) and HIV-uninfected (CORTIS-01) persons.

2.1.2.2 **Secondary Aim 2:** Compare parsimonious host-blood TB transcriptomic signatures predictive performance in HIV-infected (CORTIS-HR) and HIV-uninfected (CORTIS-01) persons.

2.1.2.3 **Secondary Aim 3:** Compare parsimonious host-blood TB transcriptomic signatures diagnostic performance, stratified by symptoms, in HIV-infected (CORTIS-HR) and HIV-uninfected (CORTIS-01) persons.

#### 2.2 STUDY ENDPOINTS

##### *2.2.1 TWO-SAMPLE ENDPOINT DEFINITION*

TB disease confirmed by positive Xpert MTB/RIF and/or MGIT culture on two or more separate sputum samples, or samples from another site if extrapulmonary disease. All aims will be evaluated using this endpoint definition.

##### *2.2.1 ONE AND TWO-SAMPLE ENDPOINT DEFINITION*

TB disease confirmed by positive Xpert MTB/RIF and/or MGIT culture on at least one sputum sample, or sample from another site if extrapulmonary disease. As exploratory analyses, all aims may also evaluated using this endpoint definition.

#### 2.3 TB DISEASE ENDPOINT ADJUDICATION ALGORITHM AND CENSORING

See CORTIS-01 and CORTIS-HR protocols and SAPs.

#### 2.4 STUDY DESIGN

See CORTIS-01 and CORTIS-HR protocols and SAPs.

#### 2.5 MEASUREMENT OF PARSIMONIOUS HOST-BLOOD TB TRANSCRIPTOMIC SIGNATURE SCORES

As part of the CORTIS-01 and CORTIS-HR studies, whole blood RNA was collected in PAXgene tubes at screening and shipped frozen to the SATVI Cape Town laboratory where RNA was extracted in a high-throughput, standardized, and reproducible fully automated procedure using a TECAN EVO Freedom robotic platform. Following cDNA-synthesis and pre-amplification steps, the parsimonious host-blood TB transcriptomic signatures listed below were run using the BioMark HD Fluidigm multiplex qRT-PCR machine and analysed using locked-down R quality control and analysis scripts:

| **Signature** | **Model*** | **Genes** | **TaqMan Assay** |
|---|---|---|---|
| Herberg2 / VIRAL2<br>(*JAMA*, 2016) | FAM89A – IFI44L | FAM89A<br>IFI44L | Custom_ARZTE3U<br>Hs00915292_m1 |
| Maertzdorf4 / DIAG4<br>(*EMBO Mol Med*, 2016) | See Suliman (*Am J Respir Crit Care Med*, 2018) | GBP1<br>IFITM3<br>P2RY14<br>ID3 | Hs00977005_m1<br>Hs03057129_s1<br>Hs01848195_s1<br>Hs00954037_g1 |
| Penn-Nicholson6 / RISK6<br>(*medRxiv*, 2019) | See Penn-Nicholson (*medRxiv*, 2019) | GBP2<br>FCGR1B<br>SERPING1<br>TUBGCP6<br>TRMT2A<br>SDR39U1 | Hs00894846_g1<br>Hs02341825_m1<br>Hs00934329_m1<br>Hs00363509_g1<br>Hs01000041_g1<br>Hs01016970_g1 |
| Roe1 / DIAG1<br>(*JCI Insight*, 2016) | BATF2 – [(TMBIM6 + CDC42 + USF2 + ACTR3)/4] | BATF2<br>Reference probes:<br>TMBIM6<br>CDC42<br>USF2<br>ACTR3 | Hs00912736_m1<br><br>Hs00162661_m1<br>Hs03044122_g1<br>Hs01100994_g1<br>Hs01029159_g1 |
| Roe3 / RISK3<br>(*Clin Infect Dis*, 2019) | [(SCARF1 + GBP5 + BATF2) / 3] – [(TMBIM6 + CDC42 + USF2 + ACTR3)/4] | SCARF1<br>GBP5<br>BATF2<br>Reference probes: | Hs01092483_m1<br>Hs00369472_m1<br>Hs00912736_m1<br>As for Roe1 |
| Suliman4 / RISK4<br>(*Am J Respir Crit Care Med*, 2018) | See Suliman (*Am J Respir Crit Care Med*, 2018) | GAS6<br>SEPT4<br>CD1C<br>BLK | Hs01090305_m1<br>Hs00910208_g1<br>Hs00957534_g1<br>Hs01017452_m1 |
| Sweeney3 / DIAG3<br>(*Lancet Respir Med*, 2016) | [(GBP5 + DUSP3)/2] − KLF2<br>See Warsinske (*JAMA Netw Open*, 2018) | GBP5<br>DUSP3<br>KLF2 | Hs00369472_m1<br>Hs01115776_m1<br>Hs00360439_g1 |
| Thompson5 / RESPONSE5<br>(*Tuberculosis (Edinb)*, 2017) | See Suliman (*Am J Respir Crit Care Med*, 2018) | UCP2<br>MAP7D3<br>STT3A<br>SMARCD3 | Hs01075224_g1<br>Hs00226257_m1<br>Hs00967491_m1<br>Hs01088251_g1 |

| | | | |
|---|---|---|---|
| | | RP11-295G20.2 | Hs01373568_m1 |

* Raw Ct values from Fluidigm Biomark qRT-PCR will be used.

The COR (RISK11) signature was run independently during the CORTIS-01 and HR studies on samples from all participants (including non-enrolled CORTIS-01 participants). The Roe1 and Roe3 signatures scores are calculated from the same Fluidigm 96.96 integrated fluidic chip (IFC) qRT-PCR runs as the RISK11 signature. The other six signatures (Herberg2, Maertzdorf4, Penn- Nicholson6, Suliman4, Sweeney3, and Thompson5) were run side-by-side on Fluidigm 192.24 IFCs, from cDNA synthesised from stored RNA aliquots of RNA extracted from all enrolled participants of CORTIS-01 and HR. For this sub-study, the performance of these eight parsimonious host-blood TB transcriptomic signatures will be compared.

#### 2.6 PROJECTED ENROLMENT, CASE ACCRUAL AND POWER

As per CORTIS-01 and CORTIS-HR protocols and SAPs.

#### 2.7 ANALYSIS POPULATIONS AND WEIGHTING

The intention to treat (ITT) and modified intention to treat population (mITT) are as defined in the CORTIS-01 and CORTIS-HR SAPs. The ITT cohort will be used for addressing primary aim 1 and secondary aim 1 (diagnostic performance in differentiating prevalent cases from combined TB negative controls and incident TB cases). The mITT cohort, which excludes prevalent TB cases, will be used for assessing signature predictive performance for risk of progression to TB disease (primary aim 2 and secondary aim 2).

##### *2.7.1* ***CORTIS-01***

Group B (COR+ observation arm) and Group C (COR- observation arm) will be included in analysis of both diagnostic and predictive performance aims, Group A (treatment efficacy arm) will be excluded from the predictive analysis, but will be included in estimating diagnostic performance at baseline (visit 2). Since the study is artificially enriched for COR+ (RISK11+) participants by design, participant-specific weights in the analysis are required to recover estimates that are applicable to the screened population, as opposed to the enrolled cohort. The COR- participants (Group C) will be up-weighted according to the empirical inverse probability of enrolling COR- participants that were screened (i.e. inverse probability weighting, IPW). Weighting will be applied as per the CORTIS-01 SAP. The 95% confidence interval will be provided for each performance metric using a non-parametric bootstrap, with sampling stratified by COR status, since the number of COR+ and COR- enrolled is fixed by the study design.

###### *2.7.1 **CORTIS-HR***

All enrolled participants will be included in analysis. There was no treatment efficacy arm in CORTIS-HR. No weighting is required, as the cohort is not enriched for COR+ participants.

### 3 STATISTICAL CONSIDERATIONS

#### 3.1 GENERAL PRINCIPLES

See CORTIS-01 and CORTIS-HR SAPs.

#### 3.2 MISSING DATA

See CORTIS-01 and CORTIS-HR SAPs.

#### 3.3 RESPONSIBILITY

Management of the clinical and laboratory data as outlined in the protocol will be the responsibility of designated laboratory technologists and doctoral scientist under supervision of Deputy Director of Immunology, SATVI and the study PI and Director SATVI.

#### 3.4 BLINDING

Participants, investigators, the data analysis team, and all members of the clinical trial team responsible for performing TB symptom and sputum screening for the purpose of endpoint determination, as well as the medical monitor, Sponsor, and data management personnel, will remain blind to participant parsimonious host-blood TB transcriptomic signature results until this SAP has been approved and signed by study sponsor. However, access to clinical and demographic data (include TB disease status) will be available to allow data cleaning and preparation of analysis script prior to database lock.

#### 3.5 DATA STORAGE

All files containing the final data will be password-protected and backed-up to the SATVI server. Data will be saved as a .csv file and compiled into one master database using R. All analysis scripts and outputs will also be backed up onto the SATVI server upon completion of the study report.

### 4 STATISTICAL METHODS

#### 4.1 SIGNATURE PERFORMANCE ANALYSIS

The performance of the signatures measured at screening will be evaluated on their ability to diagnose prevalent TB disease at baseline/enrolment and predict incident TB disease over 15-month follow up. In the CORTIS-01 and CORTIS-HR study, a pre-specified COR (RISK11) score threshold of 60% was used to differentiate correlate positive (COR+) from correlate negative (COR-) individuals. There are no pre-defined thresholds for the parsimonious host-blood TB transcriptomic signatures, thus we plan to perform receiver operating characteristic curve (ROC) analysis to select the best diagnostic and predictive signature score thresholds. Indeterminate signature score results will be excluded from primary analysis. The proportion of samples with successful signature results (i.e. those without PCR failures or indeterminate scores) will be reported. For all analyses, HIV-infected (CORTIS-HR) and HIV-uninfected (CORTIS-01) participants will be considered independently.

Score distributions for each signature in prevalent and incident TB cases, and incident-free controls will be visually represented using violin/box/scatter plots (**Figure 6.1**). Signature score distribution will be described using median and interquartile range. For CORTIS-01, we will present both weighted (section 2.7.1, above) and unweighted plots. For CORTIS-HR, plots will not be weighted (section 2.7.2, above).

Correlations between all signature scores (Spearman *rho*), as well as demographic and clinical data will be reported in a correlation matrix (**Figure 6.2**). CORTIS-01 correlations will be weighted as described in section 2.7.1 (above). For CORTIS-HR, correlations will not be weighted.

#### 4.2 PRIMARY AIM 1: SIGNATURE DIAGNOSTIC PERFORMANCE

Binary diagnostic ROC analysis will be performed separately for each signature using both CORTIS-01 and CORTIS-HR ITT cohorts (independently). These analyses will include evaluation of the area under the curve (AUC), sensitivity, specificity, positive predictive value (PPV), negative predictive value (NPV) and number needed to screen (NNS) to detect one case of TB at enrolment in HIV-infected and HIV-uninfected adults. Signature diagnostic accuracy versus the WHO target product profile (TPP) for a community-based triage or referral test to identify people suspected of having TB (World Health Organization, 2014) will be considered: For each signature, test accuracy with specificity benchmarked at 70% or sensitivity at 90% (minimum WHO triage test TPP criteria), and specificity at 80% or sensitivity 95% (optimum WHO triage test TPP criteria) will be reported.

Results will be reported in **Table 5.1** and graphically with ROC curves (**Figure 6.3**) and sensitivity/specificity versus signature score threshold plots (**Figure 6.4**).

#### 4.3 PRIMARY AIM 2: SIGNATURE PREDICTIVE PERFORMANCE

We will evaluate the changes in biomarker performance over time, including AUC, sensitivity, specificity, PPV and NPV (**Table 5.2**). We will specifically consider the following time intervals to incident TB diagnosis: 0 to 3 months, 0 to 6 months, 0 to 9 months, 0 to 12 months, and 0 to 15 months windows. We will also look at 6 month sliding windows to provide better insight into performance at different times (e.g. 0 to 6 months, 1 to 7 months, 2 to 8 months… 9 to 15 month windows).

Plots of each of these measures as a function of follow-up time will aid interpretation of the primary results at month 15 (**Figure 6.5**). Signature accuracy in predicting progression from tuberculosis infection to active disease will be compared with the WHO TPP (World Health Organization, 2017): For each signature, test accuracy with specificity benchmarked at 75% or sensitivity at 75% (minimum WHO predictive test TPP criteria), and specificity at 90% or sensitivity 90% (optimum WHO predictive test TPP criteria) will be reported.

These analyses will make use of methods and R packages developed by Zheng (*J Am Stat Assoc*, 2008), Heagerty (*Biometrics*, 2000), Pepe (*Stata J*, 2009) and others (Bansal, *Diagn Progn Res*, 2019).

#### 4.4 CONFIDENCE INTERVALS

The 95% confidence interval (95%CI) will be provided for each performance metric using a non-parametric bootstrap with 10,000 iterations. For CORTIS-01 (HIV-uninfected), bootstrap sampling will be stratified by COR status, since the number of COR+ and COR- enrolled is fixed by the study design.

#### 4.5 SECONDARY AIMS: COMPARISON OF SIGNATURE PERFORMANCE

4.5.1.1 **HIV-infected versus HIV-uninfected population signature performance:** Difference in signature performance (AUC) between HIV-infected (CORTIS-HR) and HIV-uninfected (CORTIS-01). The difference in a signature's AUC will be considered significant if the 95%CIs do not overlap. Score distributions will be visually represented with violin/box-and-scatter plots (**Figure 6.1**).

4.5.1.2 **Symptomatic versus asymptomatic population signature performance (sub-group analysis):** Difference in signature diagnostic performance (AUC) between symptomatic and asymptomatic persons will also be explored in both HIV-infected (CORTIS-HR) and HIV-uninfected (CORTIS-01) populations. The difference in a signature's AUC will be considered significant if the 95%CIs do not overlap. Score distributions will be visually represented with violin/box-and-scatter plots (**Figure 6.1**).

#### 4.6 EXPLORATORY SUB-GROUP ANALYSES

Exploratory diagnostic and predictive performance analyses may be performed by the below sub-groups. These analyses may have limited power due to small sample size and limited numbers of active TB cases. Antiretroviral (ART) status and isoniazid preventive therapy (IPT) status is determined by concomitant medication recorded during follow up and is coded as per the CORTIS-HR SAP. The difference in a performance between groups will be considered significant if the 95%CIs do not overlap. Score distributions will be visually represented with violin/box-and-scatter plots (**Figure 6.1**).

##### *4.6.1* ***CORTIS-01 AND CORTIS-HR***

4.6.1.1 Age at enrolment

4.6.1.2 Sex (male/female)

4.6.1.3 Ethnicity (Black African, Cape Mixed Ancestry)

4.6.1.4 Study site

4.6.1.5 Smoking history at enrolment (yes/no)

4.6.1.6 BMI at enrolment

4.6.1.7 Prior TB episode (yes/no)

4.6.1.8 QFT status at enrolment (<0.35, negative; ≥0.35, positive)

##### *4.6.2* ***CORTIS-HR ONLY***

4.6.2.1 ART-naïve versus ART experienced at enrolment

4.6.2.2 CD4 cell count at enrolment

4.6.2.3 Viral load at enrolment (lower than the detectable limit (<100), versus ≥ 100)

4.6.2.4 Enrolment IPT status

4.6.2.5 IPT duration during study (No IPT, IPT <6 months, IPT > 6 months)

### 5 TABLES

#### 5.1 PRIMARY AIM 1: SIGNATURE DIAGNOSTIC PERFORMANCE AT ENROLMENT (BINARY ANALYSIS) IN HIV-INFECTED (CORTIS-HR) / HIV-UNINFECTED (CORTIS-01) COHORT

| Signature | Optimal signature threshold[‡] | PCR failure or indeterminate result[†] | Sample pass rate (%) | TP | FN | TN | FP | AUC, % (95% CI) | Sensitivity[‡] % (95% CI) | Specificity[‡] % (95% CI) | PPV, % (95% CI) | NPV, % (95% CI) | NNS, n (95% CI) |
|---|---|---|---|---|---|---|---|---|---|---|---|---|---|
| RISK11 (COR) | x | x | x | x | x | x | x | x% (x–x) | x% (x–x) | 70% | x% (x–x) | x% (x–x) | x (x–x) |
| | x | x | x | x | x | x | x | | 90% | x% (x–x) | x% (x–x) | x% (x–x) | x (x–x) |
| | x | x | x | x | x | x | x | | x% (x–x) | 80% | x% (x–x) | x% (x–x) | x (x–x) |
| | x | x | x | x | x | x | x | | 95% | x% (x–x) | x% (x–x) | x% (x–x) | x (x–x) |
| Herberg2 | x | x | x | x | x | x | x | x% (x–x) | x% (x–x) | 70% | x% (x–x) | x% (x–x) | x (x–x) |
| | x | x | x | x | x | x | x | | 90% | x% (x–x) | x% (x–x) | x% (x–x) | x (x–x) |
| | x | x | x | x | x | x | x | | x% (x–x) | 80% | x% (x–x) | x% (x–x) | x (x–x) |
| | x | x | x | x | x | x | x | | 95% | x% (x–x) | x% (x–x) | x% (x–x) | x (x–x) |
| Maertzdorf4 | x | x | x | x | x | x | x | x% (x–x) | x% (x–x) | 70% | x% (x–x) | x% (x–x) | x (x–x) |
| | x | x | x | x | x | x | x | | 90% | x% (x–x) | x% (x–x) | x% (x–x) | x (x–x) |
| | x | x | x | x | x | x | x | | x% (x–x) | 80% | x% (x–x) | x% (x–x) | x (x–x) |
| | x | x | x | x | x | x | x | | 95% | x% (x–x) | x% (x–x) | x% (x–x) | x (x–x) |
| Penn-Nicholson6 | x | x | x | x | x | x | x | x% (x–x) | x% (x–x) | 70% | x% (x–x) | x% (x–x) | x (x–x) |
| | x | x | x | x | x | x | x | | 90% | x% (x–x) | x% (x–x) | x% (x–x) | x (x–x) |
| | x | x | x | x | x | x | x | | x% (x–x) | 80% | x% (x–x) | x% (x–x) | x (x–x) |
| | x | x | x | x | x | x | x | | 95% | x% (x–x) | x% (x–x) | x% (x–x) | x (x–x) |
| Roe1 | x | x | x | x | x | x | x | x% (x–x) | x% (x–x) | 70% | x% (x–x) | x% (x–x) | x (x–x) |
| | x | x | x | x | x | x | x | | 90% | x% (x–x) | x% (x–x) | x% (x–x) | x (x–x) |
| | x | x | x | x | x | x | x | | x% (x–x) | 80% | x% (x–x) | x% (x–x) | x (x–x) |
| | x | x | x | x | x | x | x | | 95% | x% (x–x) | x% (x–x) | x% (x–x) | x (x–x) |
| Roe3 | x | x | x | x | x | x | x | x% (x–x) | x% (x–x) | 70% | x% (x–x) | x% (x–x) | x (x–x) |
| | x | x | x | x | x | x | x | | 90% | x% (x–x) | x% (x–x) | x% (x–x) | x (x–x) |
| | x | x | x | x | x | x | x | | x% (x–x) | 80% | x% (x–x) | x% (x–x) | x (x–x) |

| | | | | | | | | | | | | | |
|---|---|---|---|---|---|---|---|---|---|---|---|---|---|
| | x | x | x | x | x | x | x | | 95% | x% (x–x) | x% (x–x) | x% (x–x) | x (x–x) |
| Suliman4 | x | x | x | x | x | x | x | x% (x–x) | x% (x–x) | 70% | x% (x–x) | x% (x–x) | x (x–x) |
| | x | x | x | x | x | x | x | | 90% | x% (x–x) | x% (x–x) | x% (x–x) | x (x–x) |
| | x | x | x | x | x | x | x | | x% (x–x) | 80% | x% (x–x) | x% (x–x) | x (x–x) |
| | x | x | x | x | x | x | x | | 95% | x% (x–x) | x% (x–x) | x% (x–x) | x (x–x) |
| Sweeney3 | x | x | x | x | x | x | x | x% (x–x) | x% (x–x) | 70% | x% (x–x) | x% (x–x) | x (x–x) |
| | x | x | x | x | x | x | x | | 90% | x% (x–x) | x% (x–x) | x% (x–x) | x (x–x) |
| | x | x | x | x | x | x | x | | x% (x–x) | 80% | x% (x–x) | x% (x–x) | x (x–x) |
| | x | x | x | x | x | x | x | | 95% | x% (x–x) | x% (x–x) | x% (x–x) | x (x–x) |
| Thompson5 | x | x | x | x | x | x | x | x% (x–x) | x% (x–x) | 70% | x% (x–x) | x% (x–x) | x (x–x) |
| | x | x | x | x | x | x | x | | 90% | x% (x–x) | x% (x–x) | x% (x–x) | x (x–x) |
| | x | x | x | x | x | x | x | | x% (x–x) | 80% | x% (x–x) | x% (x–x) | x (x–x) |
| | x | x | x | x | x | x | x | | 95% | x% (x–x) | x% (x–x) | x% (x–x) | x (x–x) |
| | x | x | x | x | x | x | x | | 90% | x% (x–x) | x% (x–x) | x% (x–x) | x (x–x) |

TP = True positive; FN = False negative; TN = True negative; FP = False positive; AUC = Area under the receiver operating characteristics (ROC) curve; PPV = Positive predictive value; NPV = Negative predictive value; NNS = Number needed to screen to detect 1 case of incident TB; COR = Correlate of risk.

Note: Participants who were unable to produce satisfactory sputum samples at enrolment or at end-of-study visit were assumed to be sputum negative at those time-point.

† Indeterminate results excluded. ‡ Threshold with specificity benchmarked at 70% or sensitivity at 90% (minimum WHO triage test TPP), and specificity at 80% or sensitivity 95% (optimum WHO triage test TPP).

#### 5.2 PRIMARY AIM 2: SIGNATURE PREDICTIVE PERFORMANCE FOR IDENTIFICATION OF TB DISEASE OVER A 15-MONTH PERIOD, STRATIFIED BY THE TIME INTERVAL TO DISEASE (TIME-DEPENDENT ANALYSIS) IN HIV-INFECTED (CORTIS-HR) / HIV-UNINFECTED (CORTIS-01) COHORT

Time intervals: 0 to 3 months, 0 to 6 months, 0 to 9 months, 0 to 12 months, and 0 to 15 months

1 to 6 month, 2 to 7 month, 3 to 8 month… 10 to 15 month sliding windows

| Test† and time interval to TB disease | Optimal signature threshold‡ | PCR failure or indeterminate result† | Sample pass rate (%) | Incident TB Cases | Non-incident controls | AUC, % (95% CI) | Sensitivity‡ % (95% CI) | Specificity‡ % (95% CI) | PPV‡, % (95% CI) | NPV‡, % (95% CI) |
|---|---|---|---|---|---|---|---|---|---|---|
| **Window: X to Y months** | | | | | | | | | | |
| RISK11 (COR) | x | x | x | x | x | x% (x–x) | x% (x–x) | 75% | x% (x–x) | x% (x–x) |
| | x | x | x | x | x | | 75% | x% (x–x) | x% (x–x) | x% (x–x) |
| | x | x | x | x | x | | x% (x–x) | 90% | x% (x–x) | x% (x–x) |
| | x | x | x | x | x | | 90% | x% (x–x) | x% (x–x) | x% (x–x) |
| Herberg2 | x | x | x | x | x | x% (x–x) | x% (x–x) | 75% | x% (x–x) | x% (x–x) |
| | x | x | x | x | x | | 75% | x% (x–x) | x% (x–x) | x% (x–x) |
| | x | x | x | x | x | | x% (x–x) | 90% | x% (x–x) | x% (x–x) |
| | x | x | x | x | x | | 90% | x% (x–x) | x% (x–x) | x% (x–x) |
| Maertzdorf4 | x | x | x | x | x | x% (x–x) | x% (x–x) | 75% | x% (x–x) | x% (x–x) |
| | x | x | x | x | x | | 75% | x% (x–x) | x% (x–x) | x% (x–x) |
| | x | x | x | x | x | | x% (x–x) | 90% | x% (x–x) | x% (x–x) |
| | x | x | x | x | x | | 90% | x% (x–x) | x% (x–x) | x% (x–x) |
| Penn-Nicholson6 | x | x | x | x | x | x% (x–x) | x% (x–x) | 75% | x% (x–x) | x% (x–x) |
| | x | x | x | x | x | | 75% | x% (x–x) | x% (x–x) | x% (x–x) |
| | x | x | x | x | x | | x% (x–x) | 90% | x% (x–x) | x% (x–x) |
| | x | x | x | x | x | | 90% | x% (x–x) | x% (x–x) | x% (x–x) |
| Roe1 | x | x | x | x | x | x% (x–x) | x% (x–x) | 75% | x% (x–x) | x% (x–x) |
| | x | x | x | x | x | | 75% | x% (x–x) | x% (x–x) | x% (x–x) |
| | x | x | x | x | x | | x% (x–x) | 90% | x% (x–x) | x% (x–x) |

| | | | | | | | | | | |
|---|---|---|---|---|---|---|---|---|---|---|
| | x | x | x | x | x | | 90% | x% (x–x) | x% (x–x) | x% (x–x) |
| Roe3 | x | x | x | x | x | x% (x–x) | x% (x–x) | 75% | x% (x–x) | x% (x–x) |
| | x | x | x | x | x | | 75% | x% (x–x) | x% (x–x) | x% (x–x) |
| | x | x | x | x | x | | x% (x–x) | 90% | x% (x–x) | x% (x–x) |
| | x | x | x | x | x | | 90% | x% (x–x) | x% (x–x) | x% (x–x) |
| Suliman4 | x | x | x | x | x | x% (x–x) | x% (x–x) | 75% | x% (x–x) | x% (x–x) |
| | x | x | x | x | x | | 75% | x% (x–x) | x% (x–x) | x% (x–x) |
| | x | x | x | x | x | | x% (x–x) | 90% | x% (x–x) | x% (x–x) |
| | x | x | x | x | x | | 90% | x% (x–x) | x% (x–x) | x% (x–x) |
| Sweeney3 | x | x | x | x | x | x% (x–x) | x% (x–x) | 75% | x% (x–x) | x% (x–x) |
| | x | x | x | x | x | | 75% | x% (x–x) | x% (x–x) | x% (x–x) |
| | x | x | x | x | x | | x% (x–x) | 90% | x% (x–x) | x% (x–x) |
| | x | x | x | x | x | | 90% | x% (x–x) | x% (x–x) | x% (x–x) |
| Thompson5 | x | x | x | x | x | x% (x–x) | x% (x–x) | 75% | x% (x–x) | x% (x–x) |
| | x | x | x | x | x | | 75% | x% (x–x) | x% (x–x) | x% (x–x) |
| | x | x | x | x | x | | x% (x–x) | 90% | x% (x–x) | x% (x–x) |
| | x | x | x | x | x | | 90% | x% (x–x) | x% (x–x) | x% (x–x) |

AUC = Area under the receiver operating characteristics (ROC) curve; PPV = Positive predictive value; NPV = Negative predictive value

† Indeterminate results excluded. ‡ Threshold with specificity benchmarked at 75% or sensitivity at 75% (minimum WHO TPP for test predicting progression to incident TB), and specificity at 90% or sensitivity 90% (optimum WHO TPP for test predicting progression to incident TB).

### 6 FIGURES

6.1 Test score distribution: violin/box-and-scatter plots. Plots will be weighted for CORTIS-01 (section 2.7.1) and unweighted for CORTIS-HR (section 2.7.2).

6.2 Test score correlations: correlation matrix of signature scores versus signature scores and demographic variables (spearman *rho* correlation coefficient). Plots will be weighted for CORTIS-01 (section 2.7.1) and unweighted for CORTIS-HR (section 2.7.2).

6.3 Test performance: ROC curves (sensitivity versus 100-specificity) with relevant WHO TPP criteria indicated

6.4 Test accuracy: sensitivity/specificity versus test score threshold plots (x-axis: score threshold, y-axis: sensitivity or specificity)

6.5 Time dependent analysis: AUC versus time. Sensitivity, specificity, NPV and PPV (at different thresholds) versus time, over 15-month follow-up

### 7 APPENDIX

#### 7.1 WHO DIAGNOSTIC, TRIAGE, AND PREDICTIVE TB TEST PERFORMANCE TARGET PRODUCT PROFILE

**1. Rapid biomarker-based non-sputum-based test for detecting (diagnosing) PTB (World Health Organization, 2014)**

| | Minimum diagnostic | Optimal diagnostic |
|---|---|---|
| **Sensitivity in adult PTB (overall pooled sensitivity in culture-positive cases)** | ≥65% (among both smear-positive & -negative cases) | ≥68% (among smear-negative cases only) |
| **Sensitivity in adult PTB (among smear-positive culture-positive cases only)** | >98% | ≥98% |
| **Specificity** | ≥98% | Not specified |

**2. Community-based triage or referral test to identify people suspected of having TB (World Health Organization, 2014)**

| | Minimum screening | Optimal screening |
|---|---|---|
| **Sensitivity in adult PTB (compared with confirmatory testing)** | >90% | >95% |
| **Specificity in adult PTB (compared with confirmatory testing)** | >70% | >80% |

**3. Test predicting progression from tuberculosis infection to active disease (WHO, 2017) (World Health Organization, 2017)**

| | Minimum predictive | Optimal predictive |
|---|---|---|
| **Predictive sensitivity** | ≥75% | ≥90% |
| **Predictive specificity** | ≥75% | ≥90% |

World Health Organization. 2014. High-priority target product profiles for new tuberculosis diagnostics: report of a consensus meeting. Geneva: WHO. www.who.int/tb/publications/tpp_report/en/ (Accessed 12 August 2017).

World Health Organization. 2015. The End TB Strategy: Global strategy and targets for tuberculosis prevention, care and control after 2015. Geneva: WHO. https://www.who.int/tb/strategy/End_TB_Strategy.pdf?ua=1 (Accessed September 6, 2019).

World Health Organization. 2017. Consensus Meeting Report: Development of a Target Product Profile (TPP) and a framework for evaluation for a test for predicting progression from tuberculosis infection to active disease. Geneva: WHO. http://apps.who.int/iris/handle/10665/259176 (Accessed 1 October 2017).

Zak DE, Penn-Nicholson A, Scriba TJ, Thompson E, Suliman S, Amon LM, Mahomed H, Erasmus M, Whatney W, Hussey GD, *et al.* 2016. A blood RNA signature for tuberculosis disease risk: a prospective cohort study. *Lancet.* 387 (10035):2312-2322. doi: 10.1016/S0140-6736(15)01316-1.

Zheng Y, Cai T, Pepe MS, and Levy WC. 2008. Time-dependent Predictive Values of Prognostic Biomarkers with Failure Time Outcome. *J Am Stat Assoc.* 103 (481):362-368. doi: 10.1198/016214507000001481.
